# Automation of plate inoculation and reading reduces process time in the clinical microbiology laboratory, compared to a manual workflow

**DOI:** 10.1101/2022.03.16.22272483

**Authors:** Nicole Peisach, Natalia Krotkov, Rachel Shaye, Ana María Cárdenas, Richard L. Powers

## Abstract

We evaluated the benefits of automation for clinical microbiology laboratory workflow and compared the time required for culture inoculation and plate reading between manual processes and BD Kiestra (*but* Kiestra) automation. Kiestra automation consists of different modules, each of which facilitate different tasks and work together from specimen inoculation to results reporting. We tracked the steps and measured the associated times-to-completion for both manual and automated workflows, including number of plate touches, hands-on-time (HOT), total technologist time (TTT), and walk-away time (WAT). Manual and automated processes both included 90 samples, and a total of 180 agar plates. There were three media quantity protocols used to mimic common laboratory samples: 1) 30 ‘urine’ specimens on one biplate; 2) 30 ‘urine’ specimens divided across two plates; and 3) 30 ‘blood’ or ‘wound’ specimens divided across three plates; all cultures incubated for a minimum of 18 hours prior to reading or imaging. Automation reduced HOT by 85% and overall plate touches by 88%. Automated inoculation (through implementation of the BD InoqulA module) resulted in a 100% reduction in plate touches and created 81.5-minutes of WAT. Automation of culture reading (through implementation of the BD ReadA incubation/plate imaging module and BD Synapsys Informatics) reduced HOT by 53%. Overall, laboratory automation resulted in shorter TTT and created WAT when compared to manual processes. Automation can facilitate increased processing capacity, more efficient use of the labor force, and reduced time to results in the clinical microbiology laboratory.

## INTRODUCTION

Infectious disease diagnosis often relies on the identification of bacterial pathogens in clinical specimens, such as urine, blood, and wound.^1, 2^ Traditionally, this involves manual inoculation of clinical specimens onto culture plates, which are then incubated for a period of time (typically ≥12 hours) to recover suspected bacterial pathogens. Plate reading to identify significant bacterial growth has traditionally been accomplished by experienced medical laboratory scientists. Even being a lengthy and labor-intensive process, the demand for clinical microbiology laboratory services has increased in recent years and the need for laboratory automation has become more urgent due to staffing challenges.^3-9^ In addition, manual processes are susceptible to specimen contamination, labeling error, and variability in culture quality.^10, 11^ To address these and other common problems encountered in the clinical laboratory, automation can be employed to streamline and standardize laboratory workflows. Several studies have shown improved laboratory workflow efficiency,^3-5, 10, 12-16^ result reproducibility,^4, 5, 13, 15-19^ and decreased incidence of errors ^10, 13, 18-21^ following the implementation of automation-related processes.

BD Kiestra (*but* Kiestra; Becton, Dickinson and Company; BD Life Sciences—Integrated Diagnostic Solutions, Sparks, MD) provides automation and standardization of processes associated with clinical microbiology laboratories. Kiestra is a modular system that allows each modular component to fit together in order to perform a complete laboratory workflow. BD Kiestra InoqulA specimen processor (*but* InoqulA) automates plate labeling, specimen preparation, and inoculation.^20-23^ BD Kiestra ReadA standalone (*but* ReadA), automates plate incubation, plate sorting, and high-resolution plate imaging^24^ by providing standardized digital image acquisition through the BD Synapsys Informatics Solution (*but* Synapsys) (version 3.5). Several studies have shown the benefits of implementing Kiestra automation systems in clinical microbiology laboratories, including shortening workflow time and optimizing result quality.^13, 14, 18, 19, 22, 24-26^

This report describes a step-wise utilization of automated inoculation, incubation, and imaging modules, in a simulated clinical laboratory environment, in order to streamline workflow processes. Here, we recorded each workflow step (plate setup, inoculation, and reading) and timing for plates that were processed manually, compared to the same number and types of plates that were processed via Kiestra automated modules: InoqulA (BD product number: 446973) for inoculation, ReadA (BD product number: 446948) for incubation and imaging, and Synapsys (BD product number: 444158) for digital image reading. We selected six quantifiable parameters to evaluate the efficiency for both manual and automated approaches: number of plate touches, hands-on time (HOT), setup time, cleanup time, total technologist time (TTT), and walk-away time (WAT). These metrics were chosen in order to determine whether automation can reduce the workflow burden (i.e., reducing the time required for a medical laboratory scientist to be actively hands-on in order to accomplish plate processing in the laboratory) by streamlining and automating steps, and therefore freeing up time that could be spent on other tasks that require more specialized skillsets.^7, 20-23, 27, 28^

## MATERIALS AND METHODS

### Specimens

It is typical for different clinical specimens to be plated onto various numbers of agar media, depending on the expected pathogens per source. Our protocol, utilized here, included three different media quantities per sample to mimic common clinical microbiology specimens:

1. 30 ‘urine’ specimens on one biplate (BD BBL Trypticase Soy Agar with 5% Sheep Blood (TSA II) // MacConkey II Agar biplate, Cat#221291) (10μL of specimen onto each media compartment; one plate per specimen for n=30 plates)
2. 30 ‘urine’ specimens on two plates (BD BBL Trypticase Soy Agar with 5% Sheep Blood (TSA II) plate, Cat#221261; and BD BBL MacConkey II Agar plate, Cat#221270) (10μL on each plate; 2 plates per specimen for n=60 plates)
3. 30 ‘blood’ or ‘wound’ specimens on three plates (BD BBL Trypticase Soy Agar with 5% Sheep Blood (TSA II) plate; BD BBL MacConkey II Agar plate; and BD BBL Chocolate II Agar (GC II Agar with Hemoglobin and IsoVitaleX plate, Cat#221267) (30μL on each plate; 3 plates per specimen for n=90 plates)

Manual and automated processes both used the proposed mimic specimens, therefore a total of 180 plates were testing in each method (manual and automated) for this comparison. The Chocolate plates were incubated in CO2 (5.0%) while the bi-plates, blood agar, and MacConkey II plates were incubated in ambient air (ambient O2/N2) at 35-37°C.

### Study design

#### Manual process

The manual clinical microbiology laboratory workflow was completed by a medical laboratory scientist,, identified as being proficient in this methodology and utilized independent, non-automated methods, hoods, and incubators. The medical laboratory scientist first obtained and unwrapped the media, performed specimen inoculation and then placed the inoculated plates in the incubator (either ambient O2/N2 or 5% CO2) for ≥18 hours. After retrieving the plates from the incubator, the medical laboratory scientist transported the plates to the bench top for reading, determined the presence or absence of pathogenic bacteria for each culture individually, and either marked the plates for further workup or discarded the plates.

#### Kiestra automation process

The automated microbiology laboratory workflow described in this study was completed by a medical laboratory scientist identified as being proficient in this methodology and included the InoqulA (Figure 1A) and the ReadA (Figure 1B) modules. The InoqulA module includes an automated plate sorter and an automated barcoding system for plate labeling. The medical laboratory scientist first loaded the inoculation beads and pipette tips. Next, all specimens were placed in designated InoqulA racks. The medical laboratory scientist then initialized the InoqulA system, obtained and unwrapped media plates, and loaded the plates into the automated plate sorter. Once the instrument was prepared to run and started, it then scanned the specimens, prompting the automated barcoding system to coordinate the appropriate plate types and apply the appropriate barcodes on the plates. The barcode provides traceability and coordination for subsequent imaging and reading. The InoqulA provided fully-automated liquid specimen inoculation by streaking the plates with inoculation beads.

**FIGURE 1.**
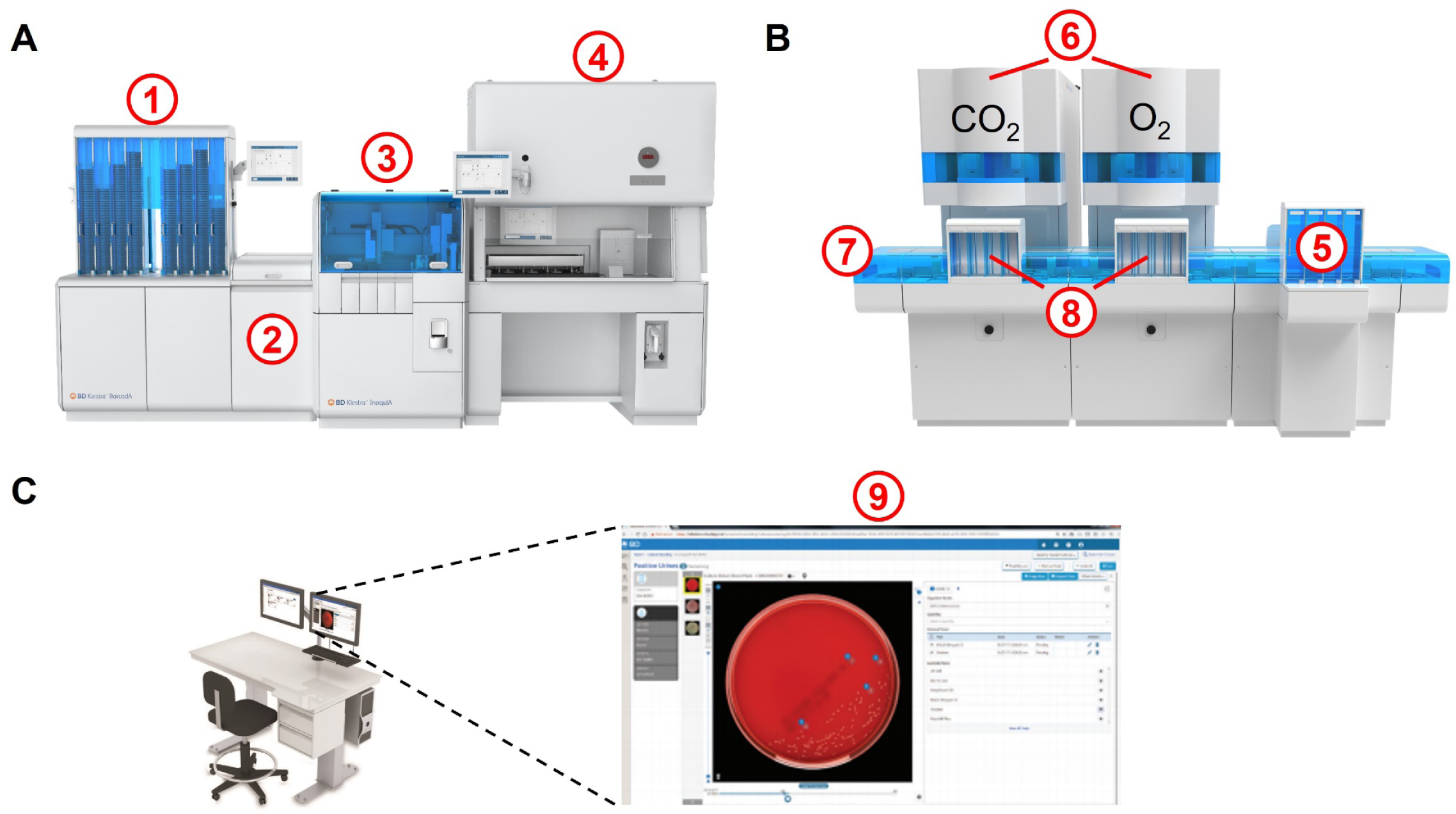
Illustrations of workflow by Kiestra modular automation. Each module can be used as standalone or together to provide different level of automation based on the laboratory needs. **(A)** An illustration of the InoqulA module. After loading specimens and plates, plates are sorted by the (1) SorterA and barcoded by the (2) BarcodA, the InoqulA (3) fully automates inoculation of liquid using rolling bead technology or (4) semi-automates inoculation of non-liquid specimens onto media plates and/or broth tubes. **(B)** An illustration of the ReadA module. Once inoculated, media plates are manually transported and loaded into the ReadA system (5) input destacker. Plates are sequentially transported to one of the two (6) ReadA modules (equipped with 5% CO2 or ambient O2/N2 incubation) via the (7) track and incubated for a pre-programmed amount of time before images are taken by the camera inside of ReadA. Once identified for further workup or disposal, the agar plates are sent to the (8) output stackers. **(C)** ReadA-obtained images are reviewed in the (9) Synapsys microbiology informatics solution, allowing a medical laboratory scientist to digitally read and determine next steps.

After inoculation and streaking by InoqulA, the medical laboratory scientist transported the plates to the ReadA (Figure 1B). The ReadA automates plate transportation, incubation, and imaging. This ReadA module includes two incubators (ambient O2/N2 and 5% CO2), which are connected to a destacker by the Kiestra track, utilized for plate transportation. After the medical laboratory scientist loaded the plates onto the destacker, the track module automatically delivered (based on barcode ID) them to the correct ReadA incubator (either O2/N2 or CO2), based on the pre-programmed incubation protocol. The built-in ReadA camera (using embedded Optis imaging technology) captured images of plates at predefined time points. The medical laboratory scientist then reviewed and analyzed digital images and determined the growth outcome from the culture plates using Synapsys software and determined whether to discard the plates or to order further workup (Figure 1C).

### Quantification parameters

This study involved a comparison of steps (Tables 1 and 2) and time-based parameters between manual and automated processes. The comparisons included the addition or elimination of workflow steps due to automation. The protocol included the number of plate touches, hands-on time (HOT) per sample, total HOT, setup time, cleanup time, total technologist time (TTT), and total walk-away time (WAT). Specifically, the number of plate touches involved the number of times when a medical laboratory scientist physically manipulated a culture plate; HOT involved the time a medical laboratory scientist was physically involved in the process and excluded setup and cleanup time; setup time involved the time a medical laboratory scientist gathered specimens, plates, and other consumables (e.g. pipette tips and inoculation beads) prior to inoculation; cleanup time involved the time spent removing used reagents and workbench clean-up; TTT involved a calculation that combined HOT, setup time, and cleanup time; WAT involved time that a medical laboratory scientist was not required to be present for a process to complete.

**Table 1.** Destacker and track time.

| **Stack pattern (No. plates)** | **Loading Destacker time** | **Destacker<sup>a</sup>** | **Cycling into ReadA time<sup>b</sup>** | **Total track time** |
| --- | --- | --- | --- | --- |
| 116 plates (30+30+30+26) | 60 s | 1020 s/17 mins | 240 s/4 mins | 21 mins |
| 120 plates (100+20) | 75 s | 240 s/4 mins | 180 s/3 mins | 7 mins |
| 60 plates (30+30) | 30 s | 500 s/8:20 mins | 160 s/2:40 mins | 11 mins |
| 60 plates (30+30) | 35 s | 389 s/6:29 mins | 141 s/2:21 mins | 8:50 mins |
| 30 plates (30) | 20 s | 375 s/6:15 mins | n/a | 6:15 mins |

<sup>a</sup>Destacker offloading to track process includes loop on track or cycle into ReadAs.
<sup>b</sup>Additional time for finishing cycling into ReadA after destacker is empty.

**Table 2.**
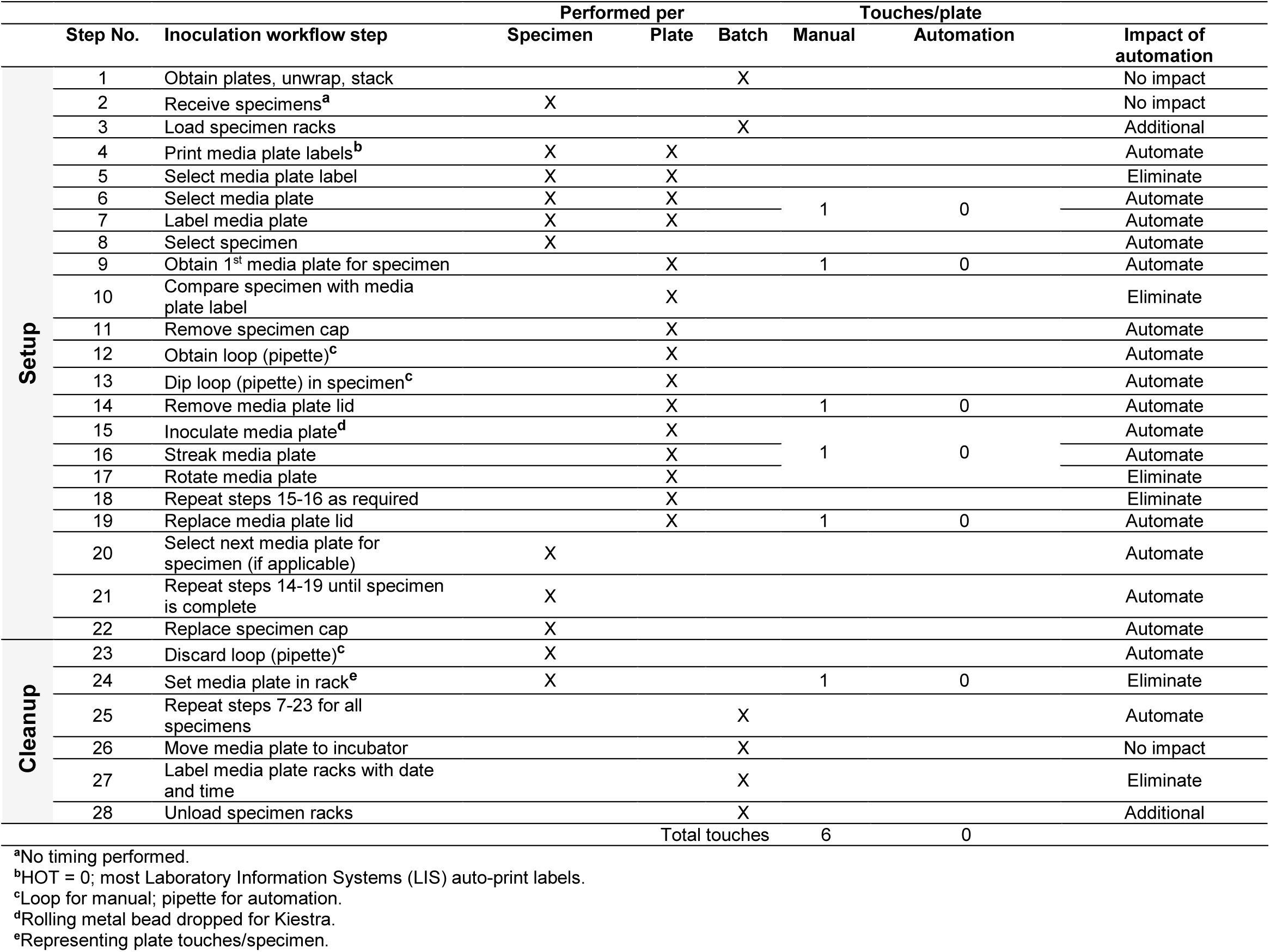
Comparison of the inoculation workflow steps between manual and automation methods.

For Kiestra automation, total track time in the ReadA system was first determined by recording the time required for loading the destacker, destacker offloading, and queue time into ReadA to establish a virtual rendering of the process for the comparison study. The time required for the destacker to offload plates onto the track varies with input plate quantity or ReadA availability (Table 1).

## RESULTS

This study compared the processing times between manual and automated processes for 90 specimens on 180 plates for each workflow process. Specimen inoculation using InoqulA resulted in the automation of 17 steps and elimination of six steps, leading to a reduction in number of plate touches from five per plate and one per specimen (with manual processes) to zero (Table 2). Once the technologist prepared the InoqulA, there was no more direct contact with the plates during the inoculation process. The technologist next handled the agar plates during their transfer from the InoqulA output stacker to the ReadA destacker. Moving inoculated plates to the incubator is the same process and approximate timing with automated and manual methods, so stack touches/moves were not counted for this step in the total plate touches for either workflow.

For incubation and media plate reading, the ReadA system either automated or eliminated six steps and resulted in two fewer touches per media plate (Table 3). Retrieving plates intended for manual reading and navigating to the correct worklist in Synapsys are considered equivalent. Therefore, setup and cleanup time differences for reading cultures via the manual method and automation are negligible.

**Table 3.** Comparison of the reading workflow steps between manual and automation methods..

| Step No. | Manual plate reading workflow step | Automation workflow step | Performed per |  |  | Touches/plate |  | Impact of automation |
| --- | --- | --- | --- | --- | --- | --- | --- | --- |
|  |  |  | Specimen | Plate | Batch | Manual | Automation |  |
| 1 | Retrieve media plates from incubators | Login to Synapsys |  |  | X |  |  | Automate |
| 2 |  | Select specimen type<sup>b</sup> |  |  | X |  |  | Additional |
| 3 |  | Access 'Ready for reading' prompt<sup>b</sup> | X |  | X |  |  | Additional |
| 4 | Select first specimen media plate set | Select first specimen from folder | X |  |  | 1 | 0 | Digital |
| 5 | Read media plates (screening) | Scan media plates |  | X |  | 1 | 0 | Digital |
| 6 | Check if 'No growth' or 'Growth' | Check if 'No growth' or 'Growth' |  |  |  |  |  | Digital |
|  | If 'No growth' (negative) | If 'No growth' (negative) | X |  |  |  |  | n/a |
| 7 | Set media plates in 'No growth' stack | Click 'Final 24' or 'Neg 24' | X |  |  |  |  | Digital |
| 8 | Select next specimen media plate set | Next specimen comes up |  |  |  |  |  | Automate |
|  | If 'Growth' (positive) | If 'Growth' (positive) |  | X |  |  |  | n/a |
| 9 | Select media plate | Select media plate |  | X |  |  |  | Digital |
| 10 | If gram negative, set in 'GN' stack<sup>c</sup> |  |  | X |  |  |  | Eliminate |
| 11 | If gram positive, set in 'GP' stack<sup>c</sup> |  |  | X |  |  |  | Eliminate |
| 12 | If mixed flora, set in 'Flora' stack<sup>c</sup> |  |  | X |  |  |  | Eliminate |
| 13 | Specify colonies<sup>a</sup> | Specify colonies |  | X |  |  |  | Digital |
| 14 | Order Maldi ID<sup>a</sup> | Order Maldi ID |  | X |  |  |  | Digital |
| 15 | Order antibiotic susceptibility test (AST) if required<sup>a</sup> | Order AST if required |  | X |  |  |  | Digital |
| 16 |  | Click 'Specimen Read'<sup>b</sup> | X |  |  |  |  | Additional |
| 17 | Select next specimen media plate set | Next specimen comes up | X |  |  |  |  | Automate |
| 18 | Repeat steps 5-19 as needed | Repeat steps 5-19 as needed |  |  |  |  |  | Digital |
| 19 | Discard negative plates<sup>a</sup> | Discard negative plates |  | X |  | 1 | 1 | No impact |
| 20 | Take positive plates to workup<sup>a</sup> | Move positive plates to next step |  | X |  |  |  | No impact |

<sup>a</sup>No timing performed<sup>b</sup>Automation only applicable<sup>c</sup>Manual mode only applicable

InoqulA workflow had longer setup times in this study, compared to the manual process, due to inoculation being performed on two different days. This is attributed to the pre-production status of the instrument and is not expected with the intended workflow of the instrument. The HOT over this time period with the InoqulA workflow was shorter, resulting in a net time reduction with automation. For all plates combined, total HOT and TTT for the manual process was 79.0 minutes and 96.0 minutes, respectively; for the automation process the times were 0 and 20.7 minutes, respectively (Table 4). In addition, the automated process yielded 81.5 minutes of WAT that were not observed with the manual process. The TTT involving all plates, combined, was 42.0 minutes for the manual process and 22.5 minutes for the automated process (Table 4).

**Table 4.** Overall time for manual and automation workflow with different protocol type.

| Protocol type | Workflow | Method | No. plates | No. Touches | HOT/plate (sec) | Total HOT (sec) | Setup time (sec) | Cleanup time (sec) | TTT (sec)<sup>a</sup> | WAT (sec)<sup>b</sup> |
| --- | --- | --- | --- | --- | --- | --- | --- | --- | --- | --- |
| One plate | Inoculation | Manual | 30 | 180 | 39 | 1170 | 90 | 60 | 1320 | 0 |
|  |  | Automation | 30 | 0 | 0 | 0 | 395 | 60 | 455 | 1280 |
|  |  | Difference<sup>c</sup> | 30 | -180 | -39 | -1170 | 305 | 0 | -865 | 1280 |
|  | Reading | Manual | 30 | 90 | 26 | 780 | 120 | 0 | 900 | n/a |
|  |  | Automation | 30 | 30 | 11 | 340 | 60 | 0 | 400 | n/a |
|  |  | Difference | 30 | -60 | -15 | -440 | -60 | 0 | -500 | n/a |
| Two plate | Inoculation | Manual | 60 | 330 | 43 | 1290 | 230 | 160 | 1680 | 0 |
|  |  | Automation | 60 | 0 | 0 | 0 | 285 | 180 | 465 | 1597 |
|  |  | Difference | 60 | -330 | -43 | -1290 | 55 | 20 | -1215 | 1597 |
|  | Reading | Manual | 60 | 180 | 14 | 420 | 0 | 0 | 420 | n/a |
|  |  | Automation | 60 | 60 | 10 | 290 | 60 | 0 | 350 | n/a |
|  |  | Difference | 60 | -120 | -4 | -130 | 60 | 0 | -70 | n/a |
| Three plate | Inoculation | Manual | 90 | 480 | 76 | 2280 | 300 | 180 | 2760 | 0 |
|  |  | Automation | 90 | 0 | 0 | 0 | 80 | 240 | 320 | 2015 |
|  |  | Difference | 90 | -480 | -76 | -2280 | -220 | 60 | -2440 | 2015 |
|  | Reading | Manual | 90 | 270 | 34 | 1020 | 0 | 180 | 1200 | n/a |
|  |  | Automation | 90 | 90 | 14 | 420 | 0 | 180 | 600 | n/a |
|  |  | Difference | 90 | -180 | -20 | -600 | 0 | 0 | -600 | n/a |
| Overall | Inoculation | Manual | 180 | 990 | 158 | 4740 (79.0 mins) | 620 (10.3 mins) | 400 (6.7 mins) | 5760 (96.0 mins) | 0 |
|  |  | Automation | 180 | 0 | 0 | 0 | 760 (12.7 mins) | 480 (8.0 mins) | 1240 (20.7 mins) | 4892 (81.5 mins) |
|  |  | Difference |  | -990 | -158 | -4740 (-79 mins) (-100%) | 140 (2.3 mins) (23%) | 80 (1.3 mins) (20%) | -4520 (-75.3 mins) (-78%) | 4892 (81.5 mins) |
|  | Reading | Manual | 180 | 540 | 74 | 2220 (37.0 mins) | 120 (2.0 mins) | 180 (3.0 mins) | 2520 (42.0 mins) | n/a |
|  |  | Automation | 180 | 180 | 35 | 1050 (17.5 mins) | 120 (2.0 mins) | 180 (3.0 mins) | 1350 (22.5 mins) | n/a |
|  |  | Difference |  | -360 | -39 (-53%) | -1170 (-19.5 mins) (-53%) | 0 (0%) | 0 (0%) | -1170 (-19.5 mins) (-46%) | n/a |
|  | Total | Manual |  | 1530 | 232 | 6960 (116 mins) | 740 (12.3 mins) | 580 (9.7 mins) | 8280 (138.0 mins) | 0 |
|  |  | Automation |  | 180 | 35 | 1050 (19.2 mins) | 880 (14.7 mins) | 660 (11 mins) | 2590 (43.2 mins) | 4892 (81.5 mins) |
|  |  | Difference |  | -1350 (-88%) | -197 (-3.3 mins) (-85%) | -5910 (-98.5 mins) (-85%) | 140 (2.3 min) (19%) | 80 (1.3 mins) (14%) | -5690 (-94.8 mins) (-69%) | 4892 (81.5 mins) |

**Abbreviations:** HOT, hands-on time; TTT, total technologist time; WAT, total walk-away time; n/a, not applicable<sup>a</sup>TTT = total HOT + setup time + cleanup time<sup>b</sup>WAT = total walk-away time (not included in workflow time)<sup>c</sup>Difference = automation- manual

For a total of 180 plates, using automation for inoculation reduced TTT by 75.3 minutes, and automation reduced reading time by 19.5 minutes. The TTT for the whole workflow was 138 minutes if using manual processes and 43.2 minutes by automation. The total differences indicate that combined automated inoculation and reading processes reduced total plate touches by 88%, largely due to the automation of inoculation steps (accounting for 990 of the 1,350 plate touches saved) (Table 4). Automation reduced the total HOT by 85%, or 98.5 minutes. Although there were slight increases in total setup and cleanup times (3.6 minutes combined), automation reduced the TTT by 69%, or 94.8 minutes (75.3 minutes for inoculation and 19.5 minutes for reading). Automation also generated 81.5 minutes of WAT, which was not present in the manual method (Figure 2).

**FIGURE 2.**
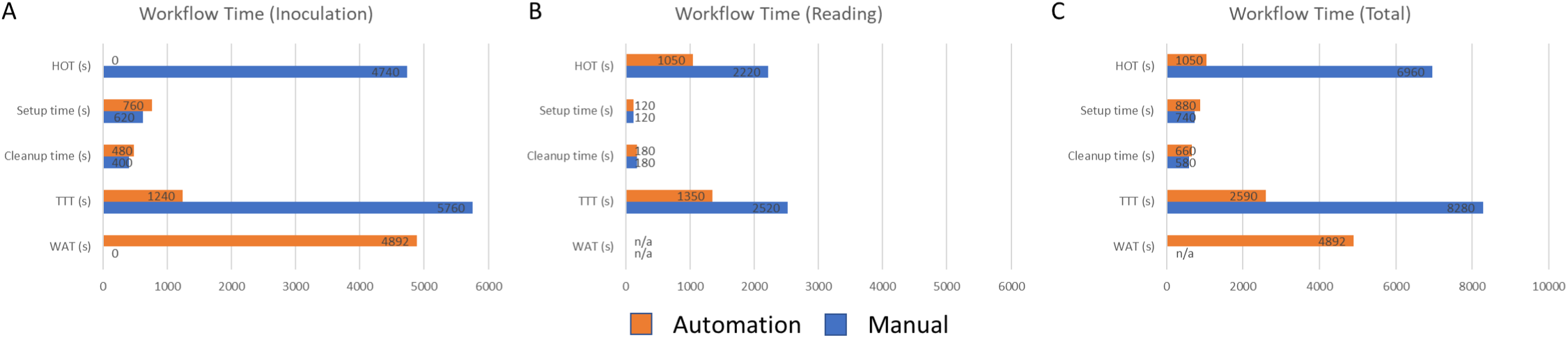
Times associated with manual and automated processes for plate inoculation and reading. (A) Workflow process times for plate inoculation are shown. Setup associated with automation showed higher setup time in this data set (see Discussion for further details) and cleanup times associated with manual and automated workflow were similar. Manual processes involved relative high HOT and TTT, with no WAT. Conversely, automated processes had no HOT, relatively low TTT, and relative high WAT. (B) Workflow process times for plate reading are shown. Setup and cleanup times associated with manual and automated processes were similar for plate reading. Manual processes involved approximately twice as much HOT, compared to automated processes, with not WAT for either workflow approach. The difference in TTT between manual and automated processes largely reflect the difference in Hot between the two approaches. (C) Total time associated with workflow processes for plate inoculation and reading, combined, for manual and automated processes. In total, automation process required about one-third of TTT compared to manual processes. WAT generated nearly doubled that for TTT with respect to automated processes whereas manual processes involved no WAT.

## DISCUSSION

Kiestra lab automation solutions offer standardized and scalable automated tools for inoculation, incubation, plate imaging, culture reading, and results reporting. However, successful implementation of laboratory automation requires thorough assessment and appropriate refinement of laboratory workflow practices that might impact system efficiency.^10, 12, 13, 29, 30^ This study provides a comprehensive evaluation of the time and steps required with a modular Kiestra-enabled automation workflow in comparison with manual processes. Utilization of the Kiestra automation system (InoqulA and ReadA) led to 88% fewer media plate touches, 85% less HOT, and 69% less TTT. In addition, automation generated 81.5 minutes of WAT during which staff could focus on other value-added tasks.

For the inoculation process, InoqulA eliminated 100% (79 minutes) of total HOT in the manual process. The only technologist time for automation was setup time and cleanup time. In this particular case, setup time and cleanup time for the automated workflow were greater (by 23% and 20%, respectively) than those same times associated with the manual workflow due to a technical event that required inoculation to occur over two days instead of one. Thus, setup and cleanup occurred twice for the automation workflow in this study compared to manual workflow, which only had one setup and cleanup period. This increase in time associated with the automation workflow is largely artificial and is not expected to apply as part of routine use in the laboratory. Plate touches during inoculation were reduced 100% (990 touches) with automation (Table 4). This is consistent with previous studies that show a significant savings in labor time and better colony recovery by automated sample preparation and plate inoculation in the microbiology diagnostic workflow.^7, 20-23, 27, 28^ The net impact of these three factors – total HOT, setup, and cleanup – was a reduction of TTT by 78% (75.3 minutes).

The ReadA system is equipped with automated imaging capacity while specimens remain in the incubator. The closed incubation setup provides optimal and stable atmosphere and temperature throughout incubation, promoting culture growth. Several studies have shown that the ReadA system yields better colony recovery and requires less time to detect growth than conventional incubators.^15, 24, 31^ The ReadA module reduced touches per plate by 67% and total HOT by 53% (19.5 minutes) during incubation and plate reading. Reading HOT per specimen was reduced on all protocol types. Because there is no WAT in the automated reading process, this same reduction of 19.5 minutes is also reflected in the TTT. With virtually no difference in setup or cleanup times, TTT was reduced by 46% (19.5 minutes). Setup time differences were negligible because the time to physically move plates from manual incubators only varied slightly (and would be lab-specific) from time-to-log in with Synapsys and navigation to the correct folders. Likewise, cleanup time differences were negligible because the time to discard negative plates or move positive plates is approximately the same in either process.

Automating inoculation, incubation, imaging, and reading can improve laboratory workflow, efficiency, and reproducibility while reducing staffing needs and artificial errors.^7, 15, 18-20, 32, 33^ By evaluating the actual time and steps required for the automation workflow, this study shows that InoqulA and ReadA modules can impact laboratory efficiency by reducing HOT and TTT, allowing lab staff to shift their attention to other value-added activities. Using InoqulA or ReadA alone to automate the inoculation or incubation and imaging process, respectively, can sufficiently save time and even generate WAT for a medical laboratory scientists to work on other valuable tasks. Additionally, the modular design can provide options for selecting some components and not others based on laboratory preference to adjust scalable automation. Overall, modular Kiestra solutions can help position microbiology laboratories to achieve more accurate and efficient testing.

### Limitations

As the system was in a controlled setting, any hardware/software issues that arose were not counted in “real-time;” therefore, any service down time as a result of mechanical or software issues were not reflected in the results, here. Since the study was conducted in a non-blinded fashion, it was not possible to eliminate outcomes bias associated with these results. Additionally, the scope of this study did not include culture workup (such as identification of AST testing) as part of the process and did not provide actual clinical microbiology results as the endpoint comparison. Therefore, this study did not capture any potential differences in diagnostic performance between automated versus manual processes. Real-world studies set in clinical laboratories might benefit from an end-to-end approach in comparing manual and automated processes. The reproducibility of results from this study will depend on the actual protocols used in the real-world clinical microbiology laboratories and other variables including medical laboratory scientist expertise involving skills that could increase or decrease manual process times compared to those reported here.

### Conclusions

Compared to manual processes, Kiestra lab automation drives reductions in total process time through automated inoculation and digital reading technology. These improvements allow for increased capacity for specimen processing and staffing in microbiology laboratories.

## Data Availability

All data produced in the present study are available upon reasonable request to the author

## Abbreviations

Total laboratory automation;

HOT: hands-on time
WAT: walk-away time
TTT: total technologist time

## ACKNOWLEDGEMENTS

The authors thank Joshua Burton and Katja Lehmann for assistance in study coordination; David Kehring, Dustin Heffron, Jon Dickerson, and Jonathan Fuchs for software engineering support; Phil Morris for maintenance, calibration, and support of laboratory instrumentation; Katja Lehmann and Harm Hoekstra for leadership support; and Yu-Chih Lin, PhD and Devin Gary, PhD for support during manuscript preparation.

